# Fear of falling and depression in older adults: the chain mediating effect of aging attitudes and social networks

**DOI:** 10.1101/2025.04.26.25326502

**Authors:** Dan Zhang, Ziqing Qi, Lulu Wu, Yali Mao, Jia Wang, Yue Zhang, Ruting Wang, Annuo Liu

**Affiliations:** School of Nursing, Anhui Medical University, No. 15 Feicui Road, Hefei 230601, Anhui Province, China

**Keywords:** Fear of falling, depression, aging attitudes, social network, chain mediation

## Abstract

**Objective:** Falls are one of the common injuries faced by older adults, and fear of falling is a common psychological stressor for older adults.This study aims to explore the relationship between fear of falling and depression in older adults and to examine the chain-mediating roles of aging attitudes and social networks.

**Method:** A stratified cluster sampling method was employed to survey 1,158 adults aged 60 and above in July–August 2022 using the Modified Falls Efficacy Scale (MFES), the Simplified Geriatric Depression Scale (GDS-15), the Attitudes to Ageing Questionnaire (AAQ), and the Lubben Social Network Scale (LSNS-6). Correlation analyses and mediation effect tests were conducted using SPSS 26.0 and SPSS PROCESS 4.2.

**Result:** The results revealed that fear of falling exerted both direct and indirect effects on depression. Aging attitudes and social networks served not only as independent mediators but also as chain mediators in the relationship between fear of falling and depression.

**Conclusion:** These findings suggest that healthcare providers should consider fear of falling as a key factor in the mental health of older adults and utilize this pathway to develop strategies aimed at reducing depression risk in this population.

## Introduction

With the aging of society, increasing attention is being paid to the physical and mental health of older adults. Depression is one of the most common mental health conditions in this population, with a global prevalence reaching as high as 35.1% among the elderly [1]. Depression contributes to adverse health outcomes and is strongly associated with reduced social functioning and lower quality of life in older adults [2–3]. According to the Circular on Comprehensively Strengthening Health Services for the Elderly issued by the National Health Commission, mental health issues such as depression and anxiety must be addressed through systematic assessment and follow-up management. Enhancing mental health in older adults is thus an essential requirement for promoting healthy aging.

Falls are a common cause of injury in the elderly and have become the leading cause of injury-related deaths among adults over 65 years of age [4]. According to Cognitive Stress Theory (CST) [5], an individual’s cognitive appraisal of stress influences their emotional responses and coping strategies. As people age, they tend to perceive a diminished ability to maintain environmental control and independent living. This, coupled with a perceived high risk of falls and reduced self-efficacy, often leads to the development of fear of falling—a major psychosocial stressor in later life. Research shows that fear of falling exists not only among those who have experienced falls but also among those who have never fallen [6]. Notably, a significant association has been observed between fear of falling and geriatric depression [7–8].

Attitudes toward aging refer to how individuals perceive and evaluate their own aging processes and old age itself [9]. Fear of falling, as a stressor, has been found to correlate significantly with aging attitudes [10]. According to Hobfoll’s Conservation of Resources Theory (COT) [11], individuals strive to acquire, retain, and protect valuable resources. The actual or anticipated loss of these resources can trigger psychological stress. With advancing age, older adults face both resource acquisition and loss. Studies have shown that a positive attitude toward aging helps older adults cope with life stressors and maintain physical functioning [12–13], whereas negative attitudes are associated with cognitive decline and a heightened risk of anxiety and depression [14–15].

A social network refers to the web of social relationships formed through interpersonal interactions [16]. According to the Social Convoy Model [17], the structure, size, and quality of an individual’s social network affect their physical and mental health throughout life. Numerous studies have shown that limited social networks contribute to depression, while strong social connections help buffer the psychological effects of negative life events and stressors [18–19].

Both aging attitudes and social networks are closely associated with depression in older adults. Research indicates that individuals’ aging attitudes influence their social relationships and access to support, which in turn affect their quality of life [20]. The Theory of Planned Behavior (TPB) posits that attitudes, subjective norms, and perceived behavioral control collectively determine behavioral intentions [21]. Thus, negative attitudes toward aging may reduce the development of social networks, increasing vulnerability to depression.

Taken together, fear of falling, aging attitudes, and social networks all impact the development of depression among older adults. However, few studies have examined the combined effects of these factors, and existing research has mostly focused on single-variable analyses. This study addresses this gap by examining fear of falling as an independent variable and exploring the mediating roles of aging attitudes and social networks in its relationship with geriatric depression. The objective is to provide a theoretical foundation for developing targeted interventions to prevent and manage depression in older adults. The hypotheses proposed are as follows. The hypothesized model is presented in Figure 1.

**Figure 1.**
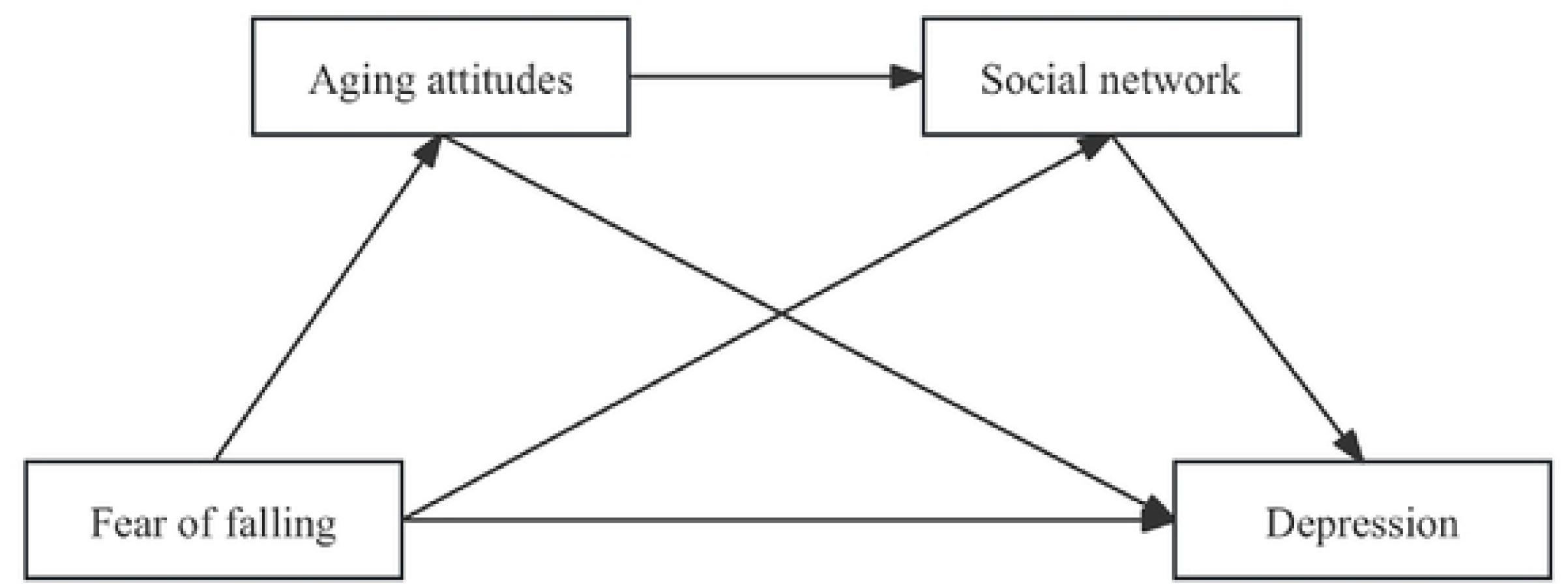
Chain mediation hypothesis model

Hypothesis 1: Fear of falling negatively predicts depression.

Hypothesis 2: Aging attitudes mediate the relationship between fear of falling and depression.

Hypothesis 3: Social networks also serve as mediators.

Hypothesis 4: Aging attitudes and social networks jointly exert a chain-mediating effect.

## 1 Materials and methods

### 1.1 Participants

A stratified cluster sampling method was adopted between July and August 2022 in Yaohai District, Hefei City. Urban and rural areas were stratified, and two streets or townships were randomly selected from each stratum. Subsequently, two communities or villages were randomly selected within each street or township to serve as survey sites. Older adults aged 60 years or above were selected as study participants. Inclusion criteria were: aged ≥60 years; cognitively sound and capable of independently completing the questionnaire; and voluntary participation with signed informed consent. Exclusion criteria included: a history of mental illness or cognitive impairment; a history of serious cardiovascular or cerebrovascular disease; and severe speech, vision, or hearing impairments. This study was approved by the Ethics Committee of Anhui Medical University (approval number: 81220209).

### 1.2 Measures

#### 1.2.1 General Information Questionnaire

A self-designed general information questionnaire was used to collect participants’ demographic characteristics. Variables included sex, age, current marital status, type of residence, education level and living status.

#### 1.2.2 Modified Falls Efficacy Scale (MFES)

The MFES, revised by Hill et al. based on the original Falls Efficacy Scale (FES) and sinicized by Hao Yanping et al. [22–23], was used to assess older adults’ confidence in avoiding falls during specific activities. The scale includes 14 items: 9 for indoor and 5 for outdoor activities. Each item is rated on an 11-point Likert scale (0 = no confidence, 5 = moderate confidence, 10 = complete confidence). The average of all items represents the final score. Lower scores indicate lower fall efficacy and higher levels of fear of falling. The Cronbach’s alpha for this scale in the present study was 0.962.

#### 1.2.3 Simplified Geriatric Depression Scale (GDS-15)

The GDS-15, translated and revised by Mei Jinrong et al. [24], contains 15 items with total scores ranging from 0 to 15. Higher scores represent more severe depressive symptoms, and a score ≥8 indicates depression. The Cronbach’s alpha for this scale in this study was 0.754.

#### 1.2.4 Attitudes to Ageing Questionnaire (AAQ)

The AAQ, revised by Huang Yifan et al. [25], assesses three dimensions: psychological growth, coping with physical change, and psychosocial loss. It contains 24 items, scored on a 5-point Likert scale (total score range: 24∼120). Items in the psychosocial loss dimension are reverse scored. Higher scores indicate more positive attitudes toward aging. The Cronbach’s alpha for the scale in this study was 0.843.

#### 1.2.5 Lubben Social Network Scale (LSNS-6)

The LSNS-6, developed by Lubben et al. [26], measures social networks using two subscales: family and friend networks, each with three items. Each item is rated on a 6-point Likert scale, with total scores ranging from 0 to 30 (0∼15 per subscale). Higher scores indicate stronger social networks. In this study, the Cronbach’s alpha coefficient was 0.776.

### 1.3 Data collection procedure

Data were collected via household visits and on-site surveys at community activity centers in July–August 2022. All investigators received uniform training. Participants were informed about the purpose of the study using standardized instructions. Upon obtaining informed consent, they were guided to complete the questionnaire on-site. Questionnaires with missing items were returned for completion. A total of 1,158 questionnaires were distributed; after excluding 39 with incomplete data, 1,117 valid responses were retained, yielding a response rate of 96.46%.

### 1.4 Statistical analysis

All data were statistically analyzed using SPSS 26.0 and PROCESS macro version 4.2. Correlation analyses were conducted, and mediation effects were tested using Model 6 in PROCESS to evaluate the sequential mediation model.

## 2 Results

### 2.1 Common method bias test

The Harman’s single-factor test was used to assess common method bias. An exploratory factor analysis was conducted on all items from the MFES, AAQ, LSNS-6, and GDS-15. Fourteen factors with eigenvalues greater than 1 were extracted, and the first factor explained 18.84% of the variance—well below the critical threshold of 40%, suggesting that common method bias was not a serious concern in this study.

### 2.2 Demographic characteristics of participants

A total of 1,117 participants were included in the analysis, comprising 440 males (39.4%) and 677 females (60.6%). Age distribution was as follows: 576 participants (51.6%) were aged 60∼69 years, 420 (37.6%) were aged 70∼79 years, and 121 (10.8%) were aged 80 years or older. Statistically significant differences in depression status were observed across age groups, education level and living status (*P* < 0.05). Detailed demographic information is presented in Table 1.

**Table 1.**
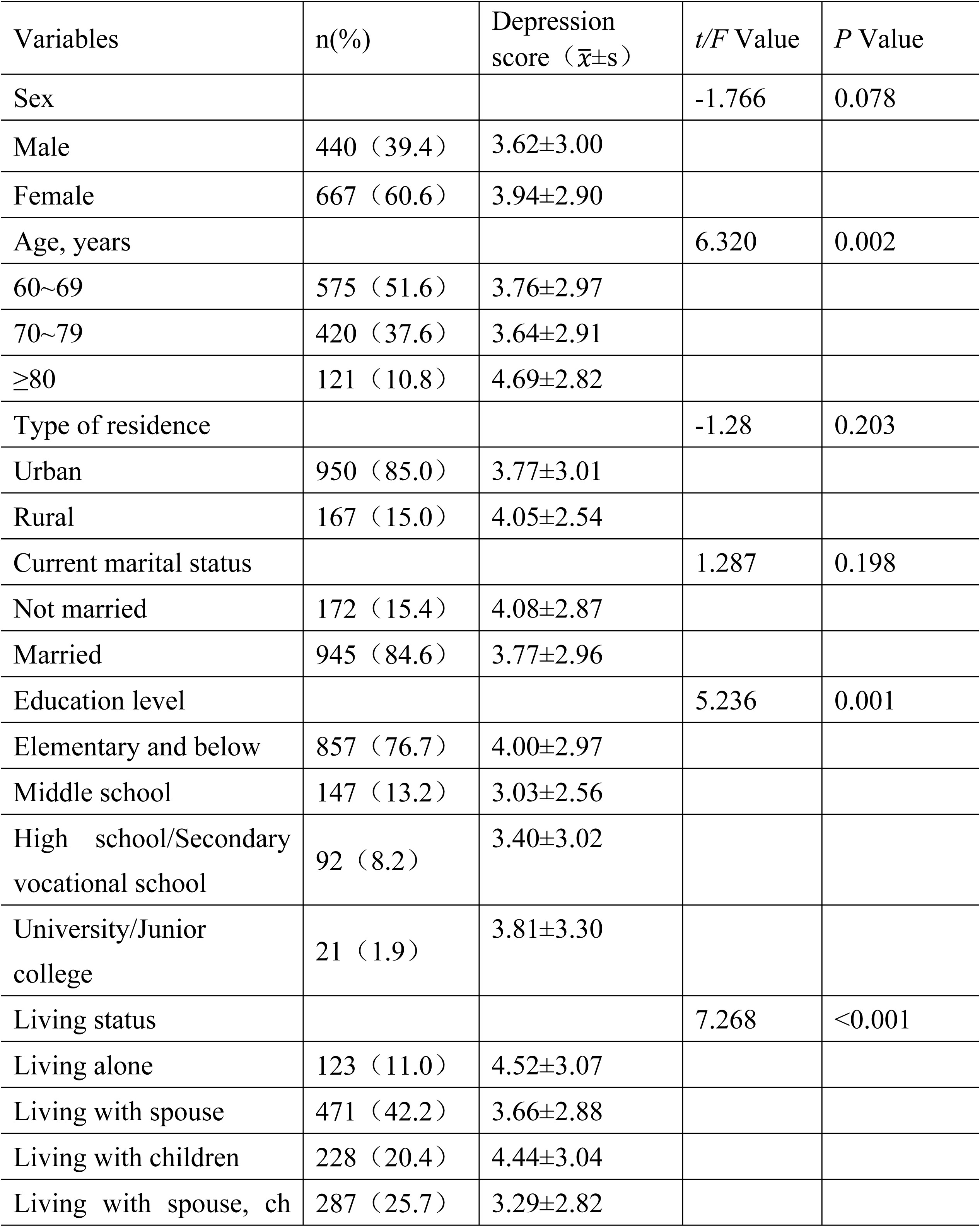
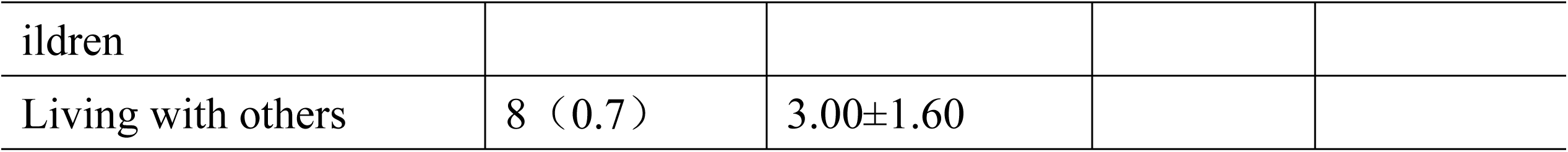
Generalization of Depression conditions in the elderly(n=1117)

### 2.3 Descriptive statistics and correlation analysis

Correlation analyses were performed on the variables and the results are shown in Table 2. There were significant correlations between fear of falling, aging attitude, social network and depression. The mean values of fear of falling, aging attitude, social network and depression scores were 9.31, 75.05, 12.28, and 3.82, respectively. fear of falling was positively correlated with aging attitude and social network (*r*=0.31, *P*<0.001; *r*=0.25,*P*<0.001), and negatively correlated with depression (*r*=-0.35, *P*<0.001). Aging attitudes were positively correlated with social networks (*r*=0.20, *P*<0.001) and negatively correlated with depression (*r*=-0.31, *P*<0.001).

**Table 2.** Descriptive statistics and correlation analysis of variables.

| variables | M±SD | 1 | 2 | 3 |
| --- | --- | --- | --- | --- |
| 1 Fear of falling | 9.31±1.39 |  |  |  |
| 2 Aging attitudes | 75.05±9.54 | 0.31*** |  |  |
| 3 Social network | 12.28±4.97 | 0.25*** | 0.20*** |  |
| 4 Depression | 3.82±2.95 | -0.35*** | -0.32*** | -0.31*** |

Note: \*\*\* indicates $P<0.001$ .

Social network was negatively correlated with depression(r=-0.31,*P*<0.001).

### 2.4 Chain mediation effect test

Using the bias-corrected percentile bootstrap method (5,000 resamples), a chain mediation model was tested with fear of falling as the independent variable, depression as the dependent variable, and aging attitudes and social networks as mediators.

The analysis results are shown in Table 3. Fear of falling significantly negatively predicted depression *(β* =-0.35, *P* < 0.001), and was positively associated with aging attitudes (*β* = 0.31, *P* < 0.001) and social networks (*β* = 0.21, *P* < 0.001). When fear of falling, aging attitudes, and social networks were simultaneously entered into the regression model predicting depression, all three emerged as significant negative predictors: fear of falling (*β* =-0.24, *P* < 0.001), aging attitudes (*β* =-0.20, *P* < 0.001), and social networks (*β* =-0.21, *P* < 0.001).

**Table 3.** Regression Analysis of Variable Relationships in Chained Mediation Models.

| Outcome Variables | Predictive variables | *R* | *R*<sup>2</sup> | *F* | $\beta$ | *t* |
| --- | --- | --- | --- | --- | --- | --- |
| Depression | Fear of falling | 0.35 | 0.12 | 158.07 | -0.35 | -12.57*** |
| Aging attitudes | Fear of falling | 0.31 | 0.10 | 117.81 | 0.31 | 10.85*** |
| Social network |  | 0.28 | 0.08 | 47.20 |  |  |
|  | Fear of falling |  |  |  | 0.21 | 6.76*** |
|  | Aging attitudes |  |  |  | 0.14 | 4.54*** |
| Depression |  | 0.46 | 0.22 | 101.49 |  |  |
|  | Fear of falling |  |  |  | -0.24 | -8.32*** |
|  | Aging attitudes |  |  |  | -0.20 | -7.21*** |
|  | Social network |  |  |  | -0.21 | -7.69*** |

Note: \*\*\* indicates $P < 0.001$ .

The mediating pathways were further examined, and the results of the total mediating effect showed (see Table 4) that aging attitudes and social networks partially mediated fear of falling and depression (Effect =-0.25, 95%CI (-0.30,-0.19)), accounting for 32.71% of the total effect. Fear of falling mainly affected depression through the following 3 mediating pathways: fear of falling indirectly affected depression through aging attitudes (Effect =-0.13, 95%CI (-0.18,-0.09)), accounting for 17.89% of the total effect; fear of falling indirectly affected depression through social networks (Effect =-0.09, 95%CI (-0.14,-0.06)), accounting for 12.42% of the total effect; and fear of falling indirectly affects depression through aging attitudes and social networks (Effect =-0.02, 95%CI (-0.03,-0.01)), accounting for 2.54% of the total effect. The diagram of the chain mediation model and the path coefficients for each variable are shown in Figure 2.

**Figure 2.**
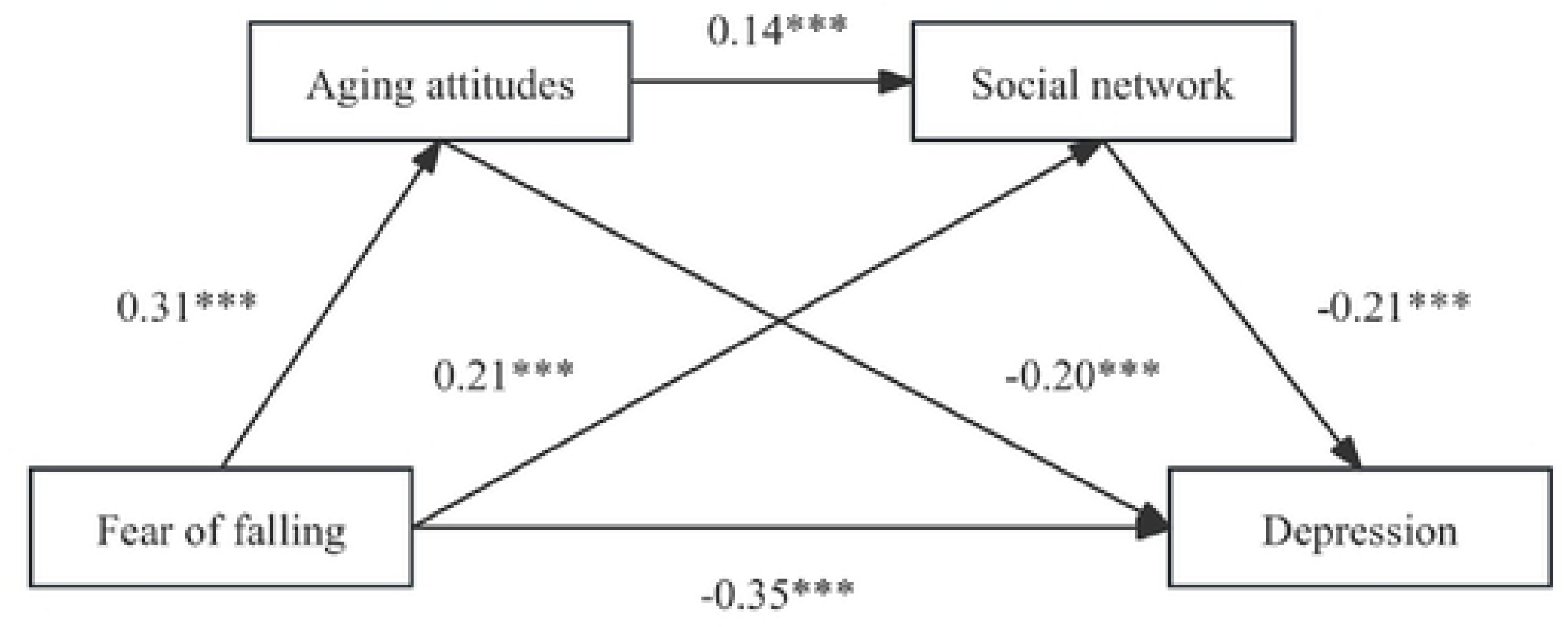
A chain-mediated model of fear of falling as a predictor of depression Note: ••• indicates p<0.00 I.

**Table 4.** Intermediary effect value and effect share.

| Type of effect | Pathway relationship | Standardized effect value | Effect proportion (%) | 95%CI |  |
| --- | --- | --- | --- | --- | --- |
|  |  |  |  | Lower | Upper |
| Direct effect | Fear of falling→Depression | -0.50 | 67.29 | -0.62 | -0.39 |
| Mediation effect | Fear of falling→Aging attitudes→Depression | -0.13 | 17.89 | -0.18 | -0.09 |
|  | Fear of falling→Social network→Depression | -0.09 | 12.42 | -0.14 | -0.06 |
|  | Fear of falling→Aging attitudes→Social network→Depression | -0.02 | 2.54 | -0.03 | -0.01 |
| Total mediation effect |  | -0.25 | 32.71 | -0.30 | -0.19 |
| Total effect |  | -0.75 | 100 |  |  |

## 3 Discussion

### 3.1 The dircet predictive effect of fear of falling on depression

The results of this study indicate that fear of falling significantly negatively predicts depression, demonstrating a dose–response relationship between the two. Specifically, older adults who experience higher levels of fear of falling are more likely to exhibit depressive symptoms. Declining physical function and the serious consequences of falls are the primary contributors to fear of falling in the elderly. One study reports that approximately 85% of older adults experience a fear of falling [27]. Impaired physical functioning, particularly gait and balance, reduces confidence in participating in social activities, leading to social withdrawal and reduced participation, which may result in social isolation [28]. Prolonged social isolation and loneliness have been shown to significantly increase the risk of depression [29]. Additionally, decreased physical and social activity can diminish muscle strength, energy, and motivation, leaving older adults in a state of physical and psychological decline [30]. Such deterioration not only lowers quality of life but also heightens the risk of developing depressive symptoms [31]. Given the reciprocal relationship between physical and mental health, it is crucial to address fear of falling as a potential determinant of mental health in older adults and to implement appropriate interventions.

### 3.2 The mediating role of aging attitudes

According to cognitive-behavioral theory [32], attitudes toward aging reflect individuals’ cognitive and emotional responses to the aging process and influence both behavior and emotional well-being. Negative attitudes toward aging have been linked to unhealthy behaviors, social withdrawal, and vulnerable personality traits, all of which contribute to psychological distress, including depression [33]. This study found a significant correlation between fear of falling and aging attitudes. Based on self-esteem protection theory [34], self-esteem serves as a psychological buffer against fear and promotes more positive attitudes toward aging [35]. Research by Bellingtier et al. further suggests that negative attitudes toward aging elevate the risk of depression by triggering maladaptive stress responses [36], whereas positive attitudes enhance coping capacity and subjective well-being, reducing the likelihood of depression [37]. The results of this study also showed that aging attitude mediated the relationship between fear of falling and depression, and the effect share (17.89%) was significantly higher than the other paths. Therefore, it is important to encourage and support older adults to adopt positive attitudes toward life and to enhance their self-confidence and self-efficacy through health education, social support, and psychological interventions to help mitigate the effects of fear of falling on depression.

### 3.3 The mediating role of social networks

Findings from the mediation analysis also reveal that social networks mediate the relationship between fear of falling and depression. As people age, their social circles tend to shrink, with family and close friends forming the core of their social networks. Research indicates that a lack of social connections can lead to social isolation, which in turn contributes to functional decline and depression [38]. Fear of falling may deter older adults from engaging in social activities, thereby reducing the size and quality of their social networks and increasing the risk of emotional distress. Social networks function as both external support systems and internal psychological anchors, buffering the negative effects of life stressors and adverse health conditions [39]. Robust family and friend networks play an essential role in helping older adults cope with the emotional consequences of fear of falling, thereby reducing the likelihood of depression [40]. Accordingly, efforts to enhance social connectivity and promote active social engagement may serve as effective interventions to alleviate the emotional impact of fear of falling.

### 3.4 Chain mediation of aging attitudes and social networks

This study confirmed a chain-mediating effect of aging attitudes and social networks in the relationship between fear of falling and depression. Positive attitudes toward aging were found to enhance social network size and quality, consistent with the findings of Menkin et al. [41], who reported that individuals with more positive aging perceptions tend to have broader social networks in later life. As a stressor, fear of falling can influence perceptions of aging. Older adults who maintain positive aging attitudes may respond to fear of falling with increased physical and social activity [11], while those with negative attitudes may withdraw from activities, leading to a contraction of their social networks and a decline in mood. Therefore, fostering positive attitudes toward aging and encouraging social interaction may help reduce the depressive effects associated with fear of falling.

### 3.5 Significance and limitations of the study

By constructing a chain mediation model between fear of falling, aging attitude, social network and depression, this study explores the correlation among the four and the psychological action mechanism of fear of falling acting on depression in older adults, providing empirical research support to further enhance aging attitude and reduce the occurrence of depression.

Fear of falling, as a common stressor in the lives of older adults, has mostly been used as a mediating or moderating variable in previous studies to examine the relationship between other variables and depression. In addition, most studies that have explored the mechanisms of depression have focused on a single level such as social networks or aging attitudes. The present study reveals how aging attitudes affect older adults’ social networks and how the two work together in relation to fear of falling and depression. This contributes to a deeper understanding of the mechanisms that shape depression in older adults. The results of this study are consistent with other studies that have highlighted the importance of aging attitudes in coping with the aging process [42]. And this study highlights the key role of social networks in maintaining the mental health of older adults. The findings suggest that we need to pay more attention to the aging attitudes of older adults, conduct regular assessment and follow-up, and encourage older adults to actively engage in social interactions and expand the size of their social networks. The findings of this study also suggest that we can strengthen the communication and cooperation among multiple disciplines, such as psychology, sociology, public health and geriatrics, in order to provide new perspectives and strategies to jointly solve the mental health problems of the elderly.

This study also has some limitations. First, this study was limited by cross-sectional research methodology, which did not allow for causal inferences. In addition, this study did not conduct a multidimensional analysis of the variables, such as distinguishing between family and friend networks in social networks, which somewhat limits the ability of health care providers to develop more specific strategies. This suggests that future studies could consider multidimensional analysis of the variables with a view to obtaining more precise findings.

## Data Availability

A confidentiality agreement was signed with the study participants before data collection for access to the data, please contact the Ethics Committee of Anhui Medical University.

## Acknowledgments

Thanks to all those who contributed to this study and to the Foundation Program for their support. This study was supported by 2023 Anhui University Natural Science Research Major Project: Research on Accurate Prediction and StageMatching Intervention of Social Isolation of Elderly Patients with Chronic Diseases Based on Digital Empowerment(2023AH040085).

## Author Contributions

Conceptualization: Dan Zhang, Annuo Liu. Data curation:Dan Zhang, Ziqing Qi, Lulu Wu.

Formal analysis: Dan Zhang, Ziqing Qi, Lulu Wu. Funding acquisition: Annuo Liu.

Investigation: Dan Zhang, Ziqing Qi, Lulu Wu,Yali Mao, Jia Wang,Yue Zhang, Ruting Wang

Methodology: Dan Zhang, Ziqing Qi, Lulu Wu,Yali Mao. Project administration: Dan Zhang,Annuo Liu.

Resources: Dan Zhang, Ziqing Qi, Lulu Wu. Software: Dan Zhang, Ziqing Qi.

Supervision: Yali Mao, Jia Wang, Yue Zhang. Validation: Jia Wang,Yue Zhang, Ruting Wang Writing – original draft: Dan Zhang.

Writing – review & editing: Dan Zhang, Annuo Liu.

